# Collapsing the list of myocardial infarction-related differential expressed genes into a diagnostic signature

**DOI:** 10.1101/2020.01.29.20019554

**Authors:** German Osmak, Natalia Baulina, Philipp Koshkin, Olga Favorova

## Abstract

Myocardial infarction (MI) is one of the most severe manifestations of coronary artery disease (CAD) and the leading cause of death from non-infectious diseases worldwide. It is known that the central component of CAD pathogenesis is a chronic vascular inflammation. However, the mechanisms underlying the changes that occur in T, B and NK lymphocytes, monocytes and other immune cells during CAD and MI are still poorly understood. One of those pathogenic mechanisms might be the dysregulation of intracellular signaling pathways in the immune cells.

In the present study we performed a transcriptome profiling in peripheral blood mononuclear cells of MI patients and controls. The machine learning algorithm was then used to search for MI-associated signatures, that could reflect the dysregulation of intracellular signaling pathways.

The genes *ADAP2, KLRC1, MIR21, PDGFD* and *CD14* were identified as the most important signatures for the classification model with L1-norm penalty function. The classifier output quality was equal to 0.911 by Receiver Operating Characteristic metric on test data. These results were validated on two independent open GEO datasets. Identified MI-associated signatures can be further assisted in MI diagnosis and/or prognosis.

Thus, our study presents a pipeline for collapsing the list of differential expressed genes, identified by high-throughput techniques, in order to define disease-associated diagnostic signatures.

## Introduction

Myocardial infarction (MI) is one of the most severe manifestations of coronary artery disease (CAD) and the leading cause of death from non-infectious diseases worldwide [1]. In most cases, MI occurs as a serious complication of atherosclerosis – a complex disease, the etiology of which is still not fully elucidated [2]. Recent studies have shown that the central component of atherosclerosis pathogenesis is a chronic vascular inflammation, resulting in endothelial dysfunction and, consequently, in an increased probability of hemodynamic abnormalities, including through the thrombosis [3]. Such a vascular lesion is emerged with the leading involvement of intimal cells (fibroblasts, endothelial and smooth muscle cells) and peripheral blood mononuclear cells (PBMC) [4]. However, the pathogenic mechanisms underlying the changes that occur in PBMC (T, B and NK lymphocytes and monocytes) during atherosclerosis and MI are still poorly understood. One of those mechanisms might be the dysregulation of intracellular signaling pathways in immune cells [5].

One way to establish MI transcriptional signatures, which include both dysregulated individual genes and signaling pathways, containing dysregulated genes, is to simultaneously study of transcriptional profiles of protein-coding genes and genes for regulatory non-coding RNA. Among non-coding RNAs, miRNAs are of particular interest in the context of robustness of biological processes, since they regulate key elements of extensive segments of signaling pathways’ networks [6–8]. To date, consistency of expressional changes of miRNAs and their target genes has been investigated in macrophages of pigs and rats with experimental MI [9] and in the whole blood of MI patients [10].

In the present study, we performed a transcriptome profiling in PBMC of MI patients and healthy individuals and revealed MI-associated signatures, consisting of individual protein-coding genes or functional patterns of genes, such as miRNA with its co-expressed target genes or combination of co-expressed genes, attributed to a definite signaling pathway.

## Materials and methods

### Pipeline

The pipeline of the study design is illustrated in Figure 1. RNA Microarray analysis was used to identify genes that were significantly (*p*<0.05) associated with MI (differentially expressed genes, DEGs). Those DEGs that have passed threshold for multiple comparisons were considered MI transcriptional signatures. Functional patterns of co-expressed DEGs were also considered MI transcriptional signatures. Such functional patterns included i) differentially expressed miRNA and its co-expressed target mRNA(s) and ii) DEGs attributed to a Reactome gene set. In the latter case, the Reactome gene sets were considered the most informative if they i) account for more than 10% of all co-expressed DEGs and/or ii) include DEGs passed multiple comparisons correction.

**Figure 1.**
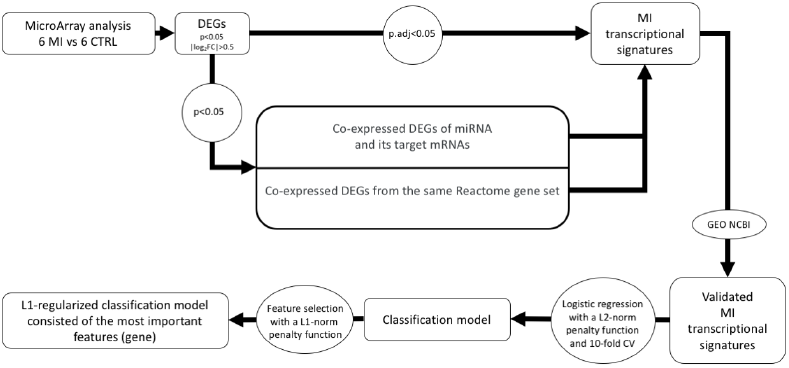
A schematic pipeline of the study for MI transcriptional signatures’ identification. DEGs – differentially expressed genes. MI – patients with myocardial infarction. CTRLs – individuals in the control group, CV - cross-validation.

The validation of identified MI transcriptional signatures was performed on two open data sets; DEGs which were not validated on at least one of these sets were excluded from further consideration. The DEGs within validated MI transcriptional signatures were used to construct binary classifiers. Given the high quality of classification and stability of the detected composite transcriptional biomarker, a logistic regression with the L1-norm penalty function was used to select the most significant DEGs on test dataset.

### Patients and controls

Six patients (all men, mean age 51.3 ± 5.9 years) with first ST-segment elevation MI were enrolled in this study. All patients were diagnosed at the National Medical Scientific Center for Cardiology (Moscow, Russia) based on symptoms of myocardial ischemia, increase of high-sensitivity cardiac troponin I (hs-cTn-I) and/or emergence of new or presumed new ST-segment elevation, new left bundle branch block or development of pathological Q waves in accordance with Third Universal Definition of MI [11]. Hs-cTn-I was measured during the initial patient assessment (from 1 to 18 h after the onset of disease symptoms). All patients underwent coronary angiography on admission and were treated according to contemporary guidelines. The characteristics of MI patients are presented in Table S1. A total of 6 CTRLs (all men, mean age 51.0 ± 7.1 years) with normal electrocardiogram, no history of CVD and diabetes mellitus were included in the study; CTRLs characteristics by smoking status and body mass index were compatible to MI patients. All participants lived in European Russia. The ethical approval was obtained from the local Ethics Committee, and written informed consent had been received from each person in accordance with the Declaration of Helsinki.

### Peripheral blood mononuclear cells collection and RNA extraction

Blood samples were collected in the morning from MI patients (24-36 h after the disease onset) and CTRLs. PBMC were isolated using Ficoll-Hypaque density gradient method (Sigma-Aldrich, St. Louis, MO, USA) within 3 h of sampling. Total RNA including small RNA was extracted using miRNeasy Mini Kit (Qiagen, Hildren, Germany) following the manufacturer’s instructions. The RNA quantity was measured using the NanoDropTM spectrophotometer (Thermo Fisher Scientific, Waltham, MA, USA); the RNA integrity was assessed by QIAxcel Advanced System (Qiagen, Hilden, Germany). Samples with RNA integrity number (RIN) value above eight were included in subsequent experiments.

### RNA Microarray analysis

The transcriptome analysis was performed using GeneChip Human Transcriptome Array 2.0, which provides the ability to analyze the expression of 44,699 protein-coding genes and 22,829 non-protein coding genes, including 1346 miRNA genes (ThermoFisher Scientific, Santa Clara, CA, USA). Briefly, total RNA (500 ng) of samples were each proceeded to poly(A)tailing and biotin ligation reactions using FlashTag Biotin HSR RNA Labeling Kit (ThermoFisher Scientific, Santa Clara, CA, USA). The biotin-labeled RNA samples were hybridized on GeneChip Human Transcriptome Array 2.0 using manufacturer’s instructions and scanned on the GeneChip Scanner 7G System. Computational analysis of the microarray data files was performed using R programming language version 3.5.1. Data processing was carried out based on the affy package written in R [12]. A biomaRt package was used to annotate the obtained data [13]. Probes demonstrating evidence for cross-hybridization, i.e. transcript sequences annotated to more than two coding genes were excluded from this study. If transcripts belong to the same gene ID, a transcript with the most detectable expression level was selected. To detect differentially expressed genes, calculate the levels of statistical significance and adjust them for multiple comparisons by Benjamini-Hochberg procedure (*p* and *padj*, respectively) the standard limma package protocol was used [14]. All expression data are deposited in the Gene Expression Omnibus international public repository under accession identification as GSE141512 [15].

### Bioinformatic analysis

MirTarBase was used to select experimentally validated target genes for miRNAs [16]. Gene set enrichment analysis (over-representation analysis) was performed using Tools of Reactome Database [17].

To construct and analyze the gene-gene interaction networks, NetworkX 2.0 package for Python was used [18]. STRING database [19] was used to find protein-protein interactions.

### Statistics analysis and Machine learning

Statistical analysis was performed using R programming language version 3.5.1. Null hypotheses were rejecting if p<0.05. To study the dependence/correlation of two continuous random variables, the Spearman’s Rank Correlation test was used. The logistic regression classifier was trained using the tools of scikit-learn v0.20.3 for Python [20]. To reduce the possible classification model overfitting the l2-norm regularization and 10-fold cross validation were used. The selection of the optimal regularization coefficient was performed by grid search using the GridSearchCV () function. The quality of the classification model was estimated by the areas under receiver operating characteristic curve (ROC-AUC). The final assessment of the quality of the classification model was carried out on the test dataset that was not used for training. For training and testing the model z-scaling of continuous features was performed at the preprocessing data stage.

### Validation analysis

The Gene Expression Omnibus database (GEO, http://www.ncbi.nlm.nih.gov/geo) was used in order to validate the obtained results. Two open data sets - GSE59867 and GSE62646 with gene expression profiles in PBMC of MI patients and healthy individuals without a history of CVD were investigated; they were obtained on GeneChip Human Gene 1.0 ST Array [transcript (gene) version]. The dataset GSE59867 included expression data of 111 MI cases and 48 CTRLs; the dataset GSE62646 - of 28 MI patients and 14 CTRLs (mixed-gender sets).

## Results

### Array-based transcriptome profiling

Transcriptome profiling in PBMC of six MI patients and six gender- and age-matched control individuals (CTRLs) was performed using GeneChip Human Transcriptome Array 2.0 (Figure 2). As a result, a total of 84 differentially expressed genes (DEGs) were identified (−0.5<Log2FC>0.5, *p*<0.05) (Table S2), from which 48 protein-coding genes and 2 miRNA genes (*MIR21* and *MIR223*) were upregulated, while 34 protein-coding genes were downregulated in MI patients. Among all observed DEGs *KLRB1* and *ADAP2* passed the threshold for multiple comparisons correction (Log2FC=-0.64, *p*.*adj*=0.0454 and Log2FC=0.64, *p*.*adj*=0.0495, respectively); both these genes were further considered as MI transcriptional signatures.

**Figure 2.**
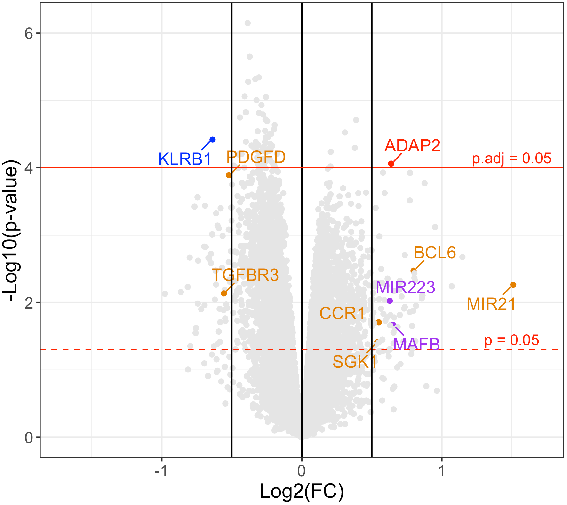
Volcano plot of gene expression changes in PBMC of MI patients compared to CTRLs. Blue dot indicates downregulated gene (Log2FC<-0.5); red dot indicates upregulated gene (Log2FC>0.5), which passed threshold for multiple comparisons (*p*.*adj*<0.05); Among differentially expressed genes (DEGs) *MIR21* and its target genes are marked in orange, *MIR223* and its target gene *−* in purple (−0.5<Log2FC>0.5, *p*<0.05).

### The search for MI transcriptional signatures: miRNA and its target mRNA(s)

Among identified DEGs (*p*<0.05) presented in Figure 2, *BCL6, CCR1, PDGFD, SGK1*, and *TGFBR3* genes were found to be targets of miR-21, while *MAFB* – target of miR-223 based on MirTarBase database. As assessed by Spearman’s correlation analysis the expression levels of *BCL6, CCR1*, and *SGK1* were positively correlated (*p*<0.001, Ro>0.9) and *PDGFD* and *TGFBR3 -* negatively correlated (*p*<0.05; Ro<-0.6) with *MIR21* expression level in MI patients and CTRLs (Figure S1). A positive correlation between the expression levels of *MAFB* and *MIR223* (*p*<0.1, Ro=0.5) was also observed (Figure S2). Thus, *MIR21* and *MIR223* genes, together with their functionally associated co-expressed target genes, were considered as two MI transcriptional signatures.

### The search for MI transcriptional signatures: Reactome gene sets

The enrichment analysis was undertaken in order to search for the functional patterns which included DEGs attributed to a Reactome gene set (Table 1). Nine Reactome gene sets were significantly overrepresented (FDR<0.05) among the 48 upregulated protein-coding genes (see above). The first three sets included each more than 10% of upregulated genes: “Immune system” *−* 22 DEGs from 2663 genes presented in the set (FDR=0.023), “Neutrophil degranulation” *−* 13 DEGs from 480 genes (FDR=0.0035) and “Cytokine Signaling in Immune system” *−* 9 DEGs from 1055 genes (FDR=0.015). “Immune system” gene set is at the highest level of the Reactome hierarchy and includes “Neutrophil degranulation” and “Cytokine Signaling in Immune system” pathways that are separately characterized by more significant overrepresentation of DEGs. So that, the DEGs from these two pathways were chosen to further analysis in the context of potential MI transcriptional signatures. Notably, “Cytokine Signaling in Immune system” pathway involves *BCL6* and *CCR1* - the target genes of miR-21, which were already included in one of the identified MI transcriptional signatures.

**Table 1.**
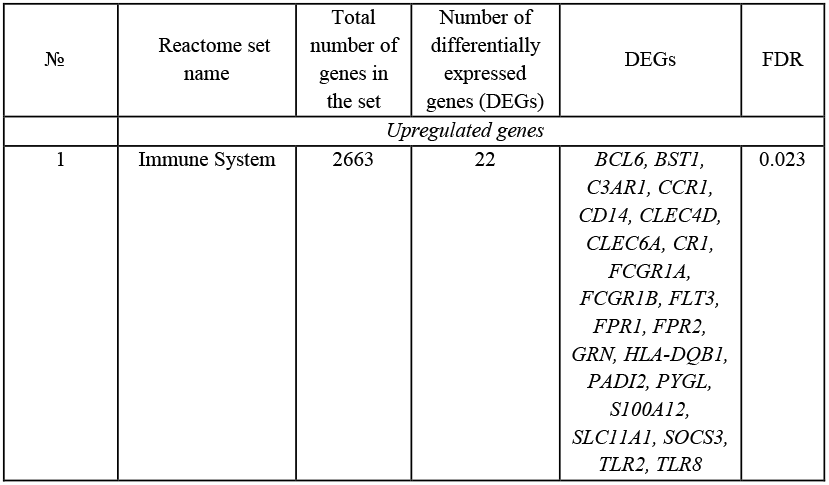
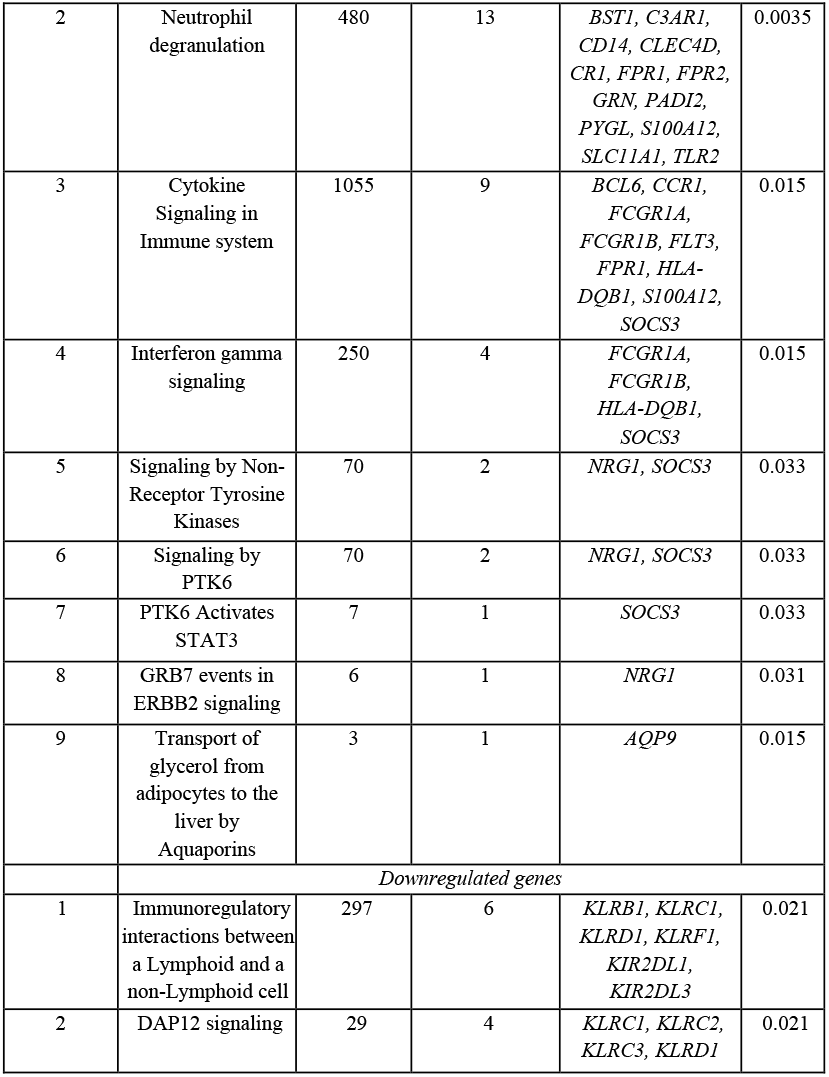
Reactome gene sets significantly overrepresented among the differentially expressed genes in PBMC from MI patients when compared to controls

As can be seen from Table 1, two Reactome gene sets were significantly overrepresented among the 34 downregulated in MI genes: “Immunoregulatory interactions between a Lymphoid and a non-Lymphoid cell” (6 DEGs from 297 genes, FDR=0.021) and “DAP12 signaling” (4 DEGs from 29 genes, FDR=0.021). Each of these sets includes more than 10% of the downregulated genes and is involved in signal transduction in lymphoid cells, namely in natural killers (NK). The “DAP12 signaling” pathway was overrepresented exclusively among the genes which encode the killer cell lectin-like receptors (KLR) expressed in NK cells, two of these DEGs (*KLRD1* and *KLRC1*) were as well observed in “Immunoregulatory interactions between a Lymphoid and a non-Lymphoid cell” pathway. The last mentioned pathway was also overrepresented by *KIR2DL1* and *KIR2DL3* genes, encoding a killer cell immunoglobulin-like receptors, the transmembrane glycoproteins expressed by NK and T cells’ subsets. Notably, *KLRB1* gene, defined previously individually as MI transcriptional signature (Log2FC=-0.64, *p*.*adj*=0.0454) was included only in “Immunoregulatory interactions between a Lymphoid and a non-Lymphoid cell” pathway. The DEGs from this pathway were chosen for further analysis in the context of potential MI transcriptional signatures.

The search for interacting genes/proteins among the DEGs from selected Reactome sets “Neutrophil degranulation”, “Cytokine Signaling in Immune system” and “Immunoregulatory interactions between a Lymphoid and a non-Lymphoid cell” (lines 2, 3 and 10 in Table 1) was performed using String database. Almost all the DEGs from the “Neutrophil degranulation” set (with the exception of the *PADI2, GRN* and *PYGL* genes) were found to interact among themselves (Figure 3A). The expression levels of these 10 interacting genes were significantly positively correlated between each other (0.93>Ro>0.51, *p*<0.05) (Figure S3). Thus, we considered the pattern of interacting genes from this pathway, namely *BST1, C3AR1, CD14, CLEC4D, CR1, FPR1, FPR2, S100A12, SLC11A1* and *TLR2* as potential MI transcriptional signature.

**Figure 3.**
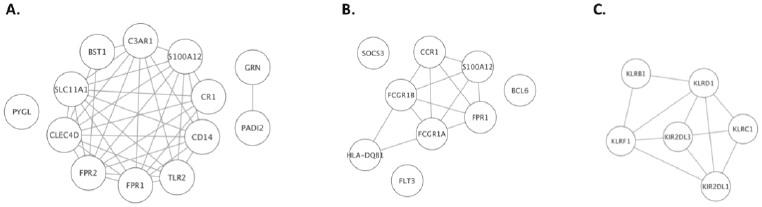
Network analysis of the Reactome gene sets “Neutrophil degranulation” (A), “Cytokine Signaling in Immune system” (B) and “Immunoregulatory interactions between a Lymphoid and a non-Lymphoid cell” (C). The edges indicate molecular interactions between nodes based on String database.

In the “Cytokine Signaling in Immune system” gene set six interacting genes (*CCR1, FCGR1A, FCGR1B, FPR1, HLA-DQB1* and *S100A12*) were found (Figure 3B), and the expression levels of these genes, with the exception of the *HLA-DQB1*, were positively correlated with each other (0.99>Ro>0.62, *p*<0.05) (Figure S4). As previously mentioned, *CCR1* is the target gene of miR-21, and has already been included to *MIR21*-containing MI transcriptional signature. According to correlation analysis, *MIR21* expression level positively correlates not only with *CCR1*, but also with *FCGR1A, FCGR1B, FPR1* and *S100A12* expression levels (0.93>Ro>0.71, *p*<0.01) (Figure S4). Thus, we considered these genes as components that extend the *MIR21*-containing MI transcriptional signature.

In Reactome pathway “Immunoregulatory interactions between a Lymphoid and a non-Lymphoid cell” different interacting gene pairs were found between all the genes from this set: *KLRB1, KLRC1, KLRD1, KLRF1, KIR2DL1, KIR2DL3* (Figure 2C) and they were predominantly characterized by significantly positive correlation between their expression levels (0.88>Ro>0.55, *p*<0.05) (Figure S5). Noteworthy, *KLRB1* gene whose differential expression passed correction for multiple comparisons and could be considered as the MI transcriptional signature, interacts with *KLRD1* and *KLRF1* from this gene set. Therefore, we included genes *KLRC1, KLRD1, KLRF1, KIR2DL1* and *KIR2DL3* in the *KLRB1*-containing MI transcriptional signature.

Overall, the conducted analysis allowed us identifying the following five MI transcriptional signatures containing all in all 29 DEGs: {*ADAP*}, {*KLRB1* + *KLRC1, KLRD1, KLRF1, KIR2DL1*, and *KIR2DL3*}, {*MIR21* + *BCL6, CCR1, PDGFD, SGK1, TGFBR3, FCGR1A, FCGR1B, FPR1*, and *S100A12*}, {*MIR223* + *MAFB*} and {*BST1, C3AR1, CD14, CLEC4D, CR1, FPR1, FPR2, S100A12, SLC11A1*, and *TLR2*}.

### The validation analysis of differential expression of genes in identified MI transcriptional signatures using GEO datasets

To confirm the differential expression of genes in identified MI transcriptional signatures we used open datasets GSE62646 and GSE59867 from GEO database, in which gene expression profiles in PBMC of MI patients and healthy individuals without a history of cardiovascular diseases (CVD) were investigated using GeneChip Human Gene 1.0 ST Array. The genes *KIR2DL1, KIR2DL3, FCGR1A* and *FCGR1B* that were according to our results included in MI transcriptional signatures were not represented on this array and were therefore excluded from the corresponding MI transcriptional signatures on further consideration. Thus, in a further analysis, 25 genes were considered.

Of the five MI transcriptional signatures we identified, the differential expression of all genes included in the *ADAP2-, KLRB1*-, and *MIR223*-containing MI transcriptional signatures was validated in both open datasets GSE62646 and GSE59867 (Table 2). The differential expression of all genes included in the *MIR21*-containing MI transcriptional signature and MI transcriptional signature from “Neutrophil degranulation” Reactome gene set was validated in GSE59867 dataset. The differential expression of a number of these genes was validated also on GSE62646 dataset with the exception of *FPR1* and *SGK1* from the *MIR21*-containing signature and *BST1, CLEC4D, FPR1, FPR2* and *TLR2* from the “Neutrophil degranulation” signature; these genes were excluded from the corresponding MI transcriptional signatures on further consideration.

**Table 2.** The expression of genes from identified MI transcriptional signatures based on our data and data obtained from GSE62646 and GSE59867 GEO datasets. Grey colour indicates p-value > 0.05.

| MI signature | Gene | Our data |  | GSE62646 |  | GSE59867 |  |
| --- | --- | --- | --- | --- | --- | --- | --- |
|  |  | logFC | p-value | logFC | p-value | logFC | p-value |
| *ADAP2*-containing<br>MI signature | *ADAP2* | 0.64 | 8.70E-05 | 0.62 | 6.40E-08 | 0.42 | 4.54E-13 |
| *KLRB1*-containing<br>MI signature | *KLRB1* | -0.64 | 3.82E-05 | -0.57 | 1.27E-03 | -0.44 | 2.85E-06 |
|  | *KLRC1* | -0.69 | 1.09E-02 | -0.80 | 3.60E-05 | -0.70 | 5.28E-10 |
|  | *KLRD1* | -0.77 | 3.78E-04 | -0.69 | 1.64E-04 | -0.70 | 7.77E-13 |
|  | *KLRF1* | -0.65 | 1.90E-03 | -0.79 | 1.06E-03 | -0.70 | 1.20E-08 |
| *MIR21*-containing<br>MI signature | *MIR21* | 1.51 | 5.45E-03 | 0.36 | 3.57E-02 | 0.80 | 2.03E-12 |
|  | *BCL6* | 0.80 | 3.38E-03 | 0.51 | 1.06E-04 | 0.48 | 2.37E-12 |
|  | *CCR1* | 0.55 | 1.97E-02 | 0.57 | 1.63E-04 | 0.67 | 1.29E-13 |
|  | *PDGFD* | -0.52 | 1.29E-04 | -0.68 | 1.56E-04 | -0.65 | 3.06E-12 |
|  | *SGK1* | 0.54 | 3.91E-02 | 0.18 | 3.00E-01 | 0.34 | 4.78E-04 |
|  | *TGFBR3* | -0.56 | 7.32E-03 | -0.59 | 1.06E-03 | -0.58 | 4.65E-11 |
|  | *FPR1* | 0.66 | 6.74E-03 | 0.28 | 5.20E-02 | 0.52 | 7.58E-12 |
|  | *S100A12* | 0.70 | 4.78E-03 | 0.43 | 4.71E-03 | 0.55 | 1.07E-10 |
| *MIR223*-containing<br>MI signature | *MIR223* | 0.63 | 9.43E-03 | 0.61 | 6.85E-05 | 0.53 | 4.85E-09 |
|  | *MAFB* | 0.65 | 2.16E-02 | 0.45 | 5.70E-04 | 0.52 | 7.80E-12 |
| Neutrophil<br>degranulation<br>MI signature | *BST1* | 0.54 | 5.13E-03 | 0.12 | 2.70E-01 | 0.50 | 9.74E-15 |
|  | *C3AR1* | 0.60 | 1.88E-02 | 0.34 | 3.14E-02 | 0.48 | 1.07E-06 |
|  | *CD14* | 0.57 | 5.09E-03 | 0.48 | 3.67E-05 | 0.60 | 5.79E-18 |
|  | *CLEC4D* | 0.81 | 3.75E-02 | -0.29 | 6.48E-02 | 0.22 | 1.72E-02 |
|  | *CR1* | 0.58 | 3.57E-02 | 0.62 | 9.67E-04 | 0.64 | 1.51E-13 |
|  | *FPR1* | 0.66 | 6.74E-03 | 0.28 | 5.20E-02 | 0.52 | 7.58E-12 |
|  | *FPR2* | 0.65 | 4.06E-02 | 0.34 | 6.67E-02 | 0.63 | 6.28E-10 |
|  | *S100A12* | 0.70 | 4.78E-03 | 0.43 | 4.71E-03 | 0.55 | 1.07E-10 |
|  | *SLC11A*<br>*1* | 0.59 | 2.38E-04 | 0.54 | 6.12E-05 | 0.50 | 2.58E-12 |
|  | *TLR2* | 0.70 | 5.09E-03 | 0.06 | 5.93E-01 | 0.22 | 3.57E-04 |

Thus, after the validation analyses MI transcriptional signatures look as follows:{*ADAP2*}, {*KLRB1* + *KLRC1, KLRD1, KLRF1*}, {*MIR21* + *BCL6, CCR1, PDGFD, TGFBR3, S100A12*}, {*MIR223* + *MAFB*} and {*C3AR1, CD14, CR1, S100A12, SLC11A1*}.

### The diagnostic value of the identified MI transcriptional signatures

The design of our study does not allow to assess the causality between MI and validated transcriptional signatures, which does not exclude the possibility of considering them as diagnostic biomarkers. Their diagnostic value can be assessed by the quality of the classification of MI patients from healthy controls. To search for such an optimal classifier, a L2 regularized logistic regression model was trained on the GSE59867 dataset (Figure 4).

**Figure 4.**
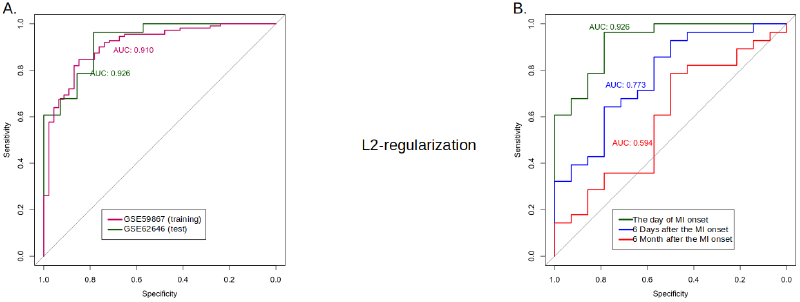
Quality and robustness of the classification model with a L2-norm penalty function based on the considered MI transcriptional signatures: {*ADAP2*}, {*KLRB1* + *KLRC1, KLRD1, KLRF1*}, {*MIR21* + *BCL6, CCR1, PDGFD, TGFBR3, S100A12*}, {*MIR223* + *MAFB*} and {*C3AR1, CD14, CR1, S100A12, SLC11A1*}. (A) Areas Under receiver operating characteristic Curve (ROC-AUC) for the training (GSE59867) and test (GSE62646) datasets. (B) Time-depended (starting from MI onset) ROC-AUC metrics of the classification model.

Figure 4A shows that MI patients at the time of admission to hospital could be classified from healthy individuals based on the selected MI transcriptional signatures, and the quality of the classification model on the test dataset (AUC=0.926) is slightly higher than on the training dataset (AUC=0.910), illustrating the stability of the model and the lack of its overfitting. While analyzing the available data from GSE62646 dataset on the levels of gene expression during the six-month follow-up after MI (Figure 4B), we observed that the classification model remains effective within 6 days after MI onset (AUC=0.773, blue line) but 6 month after MI onset the effectiveness of this model considerably decreases (AUC=0.594, red line).

For the feature selection and to reduce the number of DEGs included in the classification model we used a logistic regression with the L1-norm penalty function (Figure 5). As a result, *ADAP2, KLRC1, MIR21, PDGFD* and *CD14* genes were selected for the classification model as the most important DEGs (Figure 5A). ROC-curves constructed for these genes are demonstrated in Figures 5B and C. The comparison of ROC-curves from Figures 4 and 5 demonstrates that the quality of the classification on test dataset slightly decreased from 0.926 (dark green curve on Figure 4B) to 0.911 after applying L1 regularization (dark green curve on Figure 5B). While analyzing the changes in the quality of the classification model based on the levels of gene expression over time after MI onset, ROC-AUC values also slightly changed after applying L1 regularization (see Figures 4B and 5C). Thus, five DEGs are sufficient for the classification; among these genes DEGs from *MIR223*-containing MI transcriptional signature were not presented.

**Figure 5.**
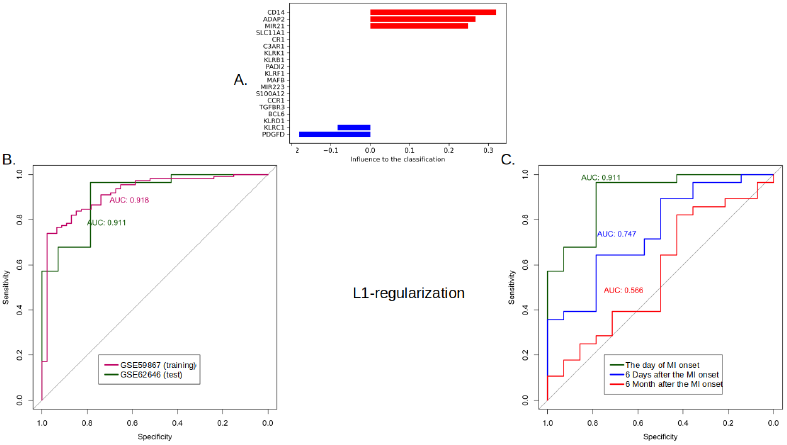
Quality and robustness of the classification model with a L1-norm penalty function based on the considered MI transcriptional signatures: {*ADAP2*}, {*KLRB1* + *KLRC1, KLRD1, KLRF1*}, {*MIR21* + *BCL6, CCR1, PDGFD, TGFBR3, S100A12*}, {*MIR223* + *MAFB*}and {*C3AR1, CD14, CR1, S100A12, SLC11A1*}. (A) Coefficients of the classification model; the most important upregulated genes *ADAP2, MIR21* and *CD14* are marked in red, downregulated genes *KLRC1* and *PDGFD* - in blue colour. (B) ROC-AUC metrics of the L1-regularized classification model consisted of *ADAP2, MIR21* and *CD14* genes. ROC-AUC were constructed using the training (GSE59867) and test (GSE62646) datasets. (C) Time-depended (starting from MI onset) ROC-AUC metrics of the L1-regularized classification model based on test dataset.

## Discussion

In the study we searched for MI transcriptional signatures (individual dysregulated genes or functional patterns of dysregulated genes) that could be potentially used in MI diagnosis. We compared the transcriptome profiles in PBMC of patients with first MI and healthy individuals using GeneChip Human Transcriptome Array 2.0 and identified five MI transcriptional signatures containing from 1 to 6 DEGs: {*ADAP2*}, {*KLRB1 + KLRC1, KLRD1, KLRF1*}, {*MIR21* + *BCL6, CCR1, PDGFD, TGFBR3, S100A12*}, *{MIR223* + *MAFB}* and {*C3AR1, CD14, CR1, S100A12, SLC11A1*}.

In order to select the most important for classification DEGs, further research steps included: validation on open datasets GSE62646 [21] and GSE59867 [22], construction of L2 regularized logistic regression model for estimation the diagnostic value of the MI transcriptional signatures and feature selection using L1-norm penalty function. This approach allowed to exclude from the classification model MI transcriptional signature (*MIR223* + *MAFB*) as insufficiently effective and to reduce the number of DEGs from other signatures to *ADAP2, KLRC1, MIR21, PDGFD* and *CD14*. According to the ROC-AUC analysis the obtained classification model, including 5 genes, is enable classifying MI patients and healthy controls with a quality of 0.911 while the quality of initial classification model, including 18 genes, was equal to 0.926. Thus, a decrease in the number of genes did not significantly affect the quality of the model. A comparable decline in the quality of both classification models over time from MI onset was shown; this decline occurs rather slowly, for days and weeks.

Consider consistently the characteristics of genes-classifiers. The gene *ADAP2* encodes ArfGAP With Dual PH Domains 2 protein and was designated in our study as individual MI transcriptional signature; no data on the involvement of this gene in the development of CVD and/or its complications were found. However, the product of this gene was shown to be involved in heart development, and its dysfunction presumably is associated with cardiovascular malformations in NF1 microdeletion syndrome [23].

The gene *KLRC1* refers to a MI transcriptional signature containing the genes of killer cell lectin-like receptors (KLR) that encode a family of transmembrane proteins, characterized by a type II membrane orientation and the presence of a C-type lectin domain; they are predominantly expressed in NK cells. The association of some genes from this signature (*KLRD1* and *KLRC1*) with MI or its complications was previously shown by Maciejak et al and Kiliszek et al, whose data were used for validation analysis in our study [21,22]. We have shown that the expression of the genes *KLRB1, KLRC1, KLRD1, KLRF1* is consistently decreased in MI, that is in a good accordance with the study by Yan et al [24], where a loss of NK cell activity was found in patients with acute MI, in particular, due to a decrease in *KLRB1* expression.

The *MIR21* gene and target genes of miR-21 were included in one MI transcriptional signature, composed mainly of genes from “Cytokine Signaling in Immune system” pathway. Thus, in addition to cytokine signaling pathways, which role in MI development was previously described [25], we have identified and validated on independent GEO datasets the influence of miR-21 through the regulation of this pathway in PBMC during MI. The functional role of miR-21 in cardiac tissue has been studied for a long time, and by now a large amount of data has been accumulated on this subject [26], while in PBMC its role remains unclear. In one of the studies the negative correlation of miR-21 expression level in MI with the levels of IL-1β, IL-6, and TNF-α cytokines was shown due to regulatory effect of this miRNA on the expression of *KBTBD7*; this gene encodes a member of BTB-kelch proteins, kelch repeat and BTB (POZ) domain containing 7, which promotes inflammatory responses in macrophages [27]. In turn, in our study, the association of miR-21 and its target genes *PDGFD, TGFBR3, CCR1* and *BCL6* expression levels with MI was demonstrated, from which *PDGFD* gene encoded platelet derived growth factor D was found to be the most important based on the results of L1 regularization. The genes of the PDGF family and their involvement in the pathogenesis of various diseases are well studied; in particular, PDGFD is known to be involved in the fibrosis and neovascularization of the cardiac tissue [28].

The gene *CD14* encodes a receptor on the surface of myeloid cells, which participates in CD14/TLR4/MD2 signaling pathway involved in the recognition of lipopolysaccharides [29]. This gene was identified in our study as a component of “Neutrophil degranulation” pathway. The neutrophils are known to be actively involved in the development and elimination of MI consequences [30]. Furthermore, polymorphic variants in *CD14* gene were found to be associated with MI [31].

Further investigations are implicitly needed to clarify the causality between MI and the identified MI-associated signatures.

The data on differential expression of a number of genes in PBMC of MI patients obtained in our study were validated on two independent datasets that indicates their value. The identified DEGs could be suitable for the prediction of the first MI before the appearance of the disease symptoms, as it was previously described for some miRNAs [32]. Further investigations are implicitly needed to clarify the functional role of the identified MI-associated genes in the development of this disease.

## Conclusions

The present study implements the pipeline designed to the collapsing the list of differential expressed in MI genes into a diagnostic signature; the obtained classification model is enable classifying MI patients and healthy controls with a quality of 0.911 on a test data. This pipeline could be useful in high-throughput data analysis for the searching of diagnostic signature of other diseases.

## Data Availability

https://www.ncbi.nlm.nih.gov/geo/query/acc.cgi?acc=GSE141512

## Authors’ contributions

Conceptualization, G.O., N.B. and O.F.; Data curation, G.O. and P.K.; Formal analysis, G.O.; Investigation, G.O., N.B. and P.K.; Methodology, N.B.; Project administration, O.F.; Validation, G.O.; Visualization, G.O.; Writing – original draft, G.O. and O.F.; Writing – review & editing, G.O., N.B. and O.F‥

## Acknowledgments

We thank Dr. Roman Shakhnovich, Dr. Nino Kulava and PhD Natalia Matveeva from National Medical Research Center for Cardiology, Russia, for their assistance with MI patients and controls collection. We appreciate PhD Alexander Favorov for valuable advices while producing the manuscript.

---

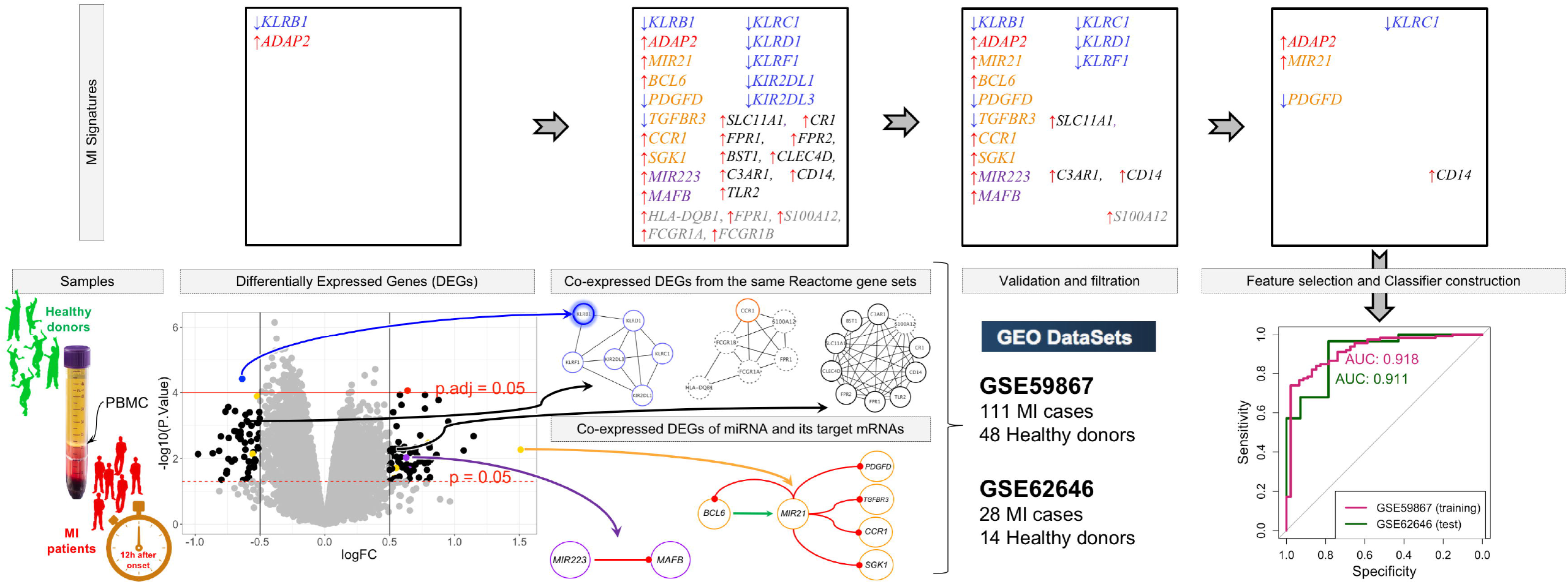

